# Statistical modeling for quality risk assessment of clinical trials: follow-up at the era of remote auditing

**DOI:** 10.1101/2021.07.12.21260214

**Authors:** Björn Koneswarakantha, Timothé Ménard

## Abstract

**Background:** As investigator site audits have largely been conducted remotely during the COVID-19 pandemic, remote quality monitoring has gained some momentum. To further facilitate the conduct of remote Quality Assurance (QA) activities for clinical trials, we developed new quality indicators, building on a previously published statistical modeling methodology.

**Methods:** We modeled the risk of having an audit or inspection finding using historical audits and inspections data from 2011 - 2019. We used logistic regression to model finding risk for 4 clinical impact factor (CIF) categories: Safety Reporting, Data Integrity, Consent and Protecting Endpoints.

**Results:** We could identify 15 interpretable factors influencing audit finding risk of 4 out of 5 CIF categories. They can be used to realistically predict differences in risk between 25 and 43% for different sites which suffice to rank sites by audit and inspection finding risk.

**Conclusion:** Continuous surveillance of the identified risk factors and resulting risk estimates could be used to complement remote QA strategies for clinical trials and help to manage audit targets and audit focus also in post-pandemic times.

## Background

During the COVID-19 pandemic investigator site audits have largely been conducted remotely relying on digital technology. Site-specific quality indicators could be used to identify quality risk of specific sites. To validate potential quality indicators, we could estimate their influence on the probability of past audit findings. We have already shown that such an approach was feasible in the Good Clinical and Pharmacovigilance Practices areas and that we could overcome challenges such as low signal-to-noise ratios and relatively small data sets with high ratios of missing values [1,2]. Having expanded our operational dataset, we have gathered historic investigator site audits/inspection findings from 2011 - 2019 and grouped them into 5 clinical impact factors (CIFs). We then generated a large set of operational features for each audited or inspected site and used them to model the probability of having a finding in a given category. Our goal was to obtain a set of interpretable features which contribute to fairly accurate risk estimates rather than building a classification model for audit findings. In total we could identify 13 site-specific quality indicators that are clearly associated with audit and inspection finding risk for 4 different CIFs. This was a major improvement over our last modeling iteration in which we could only identify 5 site-specific quality indicators. This improvement is mostly attributable to the additional operational site data which allowed us to create new modeling features that were previously unavailable.

## Methods

Audit finding, inspection and all clinical trial data were gathered from Roche internal data sources and contained (808 audits/inspections over 9 years). A full description can be found in the code repository. The risk for having one or more audit and inspection findings in a given category was modeled using logistic regression as previously described [1].

### Data Preparation and Feature Selection

Operational features (based on Adverse Events (AE), Issues and Deviations and Data Queries) were engineered to reflect the state of the site (at the time of the audit/inspection). They were complemented by site characteristics such as geographic population and study characteristics such as therapeutic area. To account for the number of patients and the individual study progress of each patient, we normalized features by either number of patient visits or total number of days that have passed for all patients since enrollment (referenced as days on study). As critical thresholds for quality indicators can be protocol specific, we also calculated the study percent rank for some features (e.g. percentage of missed visits and number of parallel trials) which indicated the percentage of sites in the same study that had a lower value.

To account for non-linear relationships between features and risk, continuous features were normalized using a Yeo Johnson power transformation [3] and binned into 5 groups with the same value range. For missing values all resulting binary bins were set to zero. Finding frequencies for each bin and categorical features were examined and promising candidates were preselected. The set of preselected features was narrowed down by fitting logistic regression models to a training data subset (2011-2015). We then iteratively removed uninterpretable features based on Subject Matter Expert (SME) review, and merged bins to facilitate their interpretation, to obtain a final feature set which was used to fit a model on the entire data set. We checked for multicollinearity between the features of each model by calculating the variance inflation factor (VIF) for each coefficient, and found that the maximum VIF per model was never greater than 2 (Table 1), which is well below the accepted critical threshold of 5 [4]. Sites that had missing values for all final features were removed before model fitting, otherwise for missing values all derived bins from that feature were set to zero.

**Table 1.** Mean Modelling Performance per CIF Model - Mean AUC and Brier Score Including Standard Error (SE) were Calculated Based on Test Set Predictions Derived from Time Series Cross-Validation Strategy with One Value per Year from 2011 - 2018

| Clinical Impact Factor | Maximum VIF | Mean AUC ±SE | Mean Brier Score ±SE | Calibrated Prediction Range % | Base Rate Probability % |
| --- | --- | --- | --- | --- | --- |
| Consent | 1.25 | 0.61 ± 0.15 | 0.24 ± 0.01 | 34-61( $\Delta$ 27) | 46 |
| Data Integrity | 1.11 | 0.60 ± 0.1 | 0.19 ± 0.02 | 49-85( $\Delta$ 36) | 73 |
| Protecting Endpoints | 1.05 | 0.59 ± 0.06 | 0.23 ± 0.01 | 54-79( $\Delta$ 25) | 69 |
| Safety | 1.97 | 0.63 ± 0.7 | 0.25 ± 0.01 | 26-69 ( $\Delta$ 43) | 47 |
| Sponsor Oversight | 1.05 | 0.53 ± 0.06 | 0.24 ± 0.01 | 63-65 ( $\Delta$ 2) | 64 |

### Model Validation

To validate the models and to get a performance estimate, we used time series cross validation [5] in which data from each year would be used as a test set for a model fit with all data from previous years (see Fig. 1). The receiver operator characteristics area under the curve (AUC) and Brier Scores were calculated for each cross validation test set and mean and standard error were calculated. As we were more interested in accurate risk estimates than in classification, we proceeded to fit a calibration model. The probability risk estimates of the test set predictions were divided into 4 bins of semi-equal range with a minimum of 100 test predictions per bin. For each bin, the predicted risk was averaged and the actual observed risk was calculated. To fit the calibration model, we performed a linear regression on the mean predicted risk versus the observed risk. In order to avoid extreme predictions by combinations of risk factors that are unvalidated, we calculated a lower and upper range limit using the average observed risk of the 200 lowest and 200 highest risk estimates.

**Figure 1.**
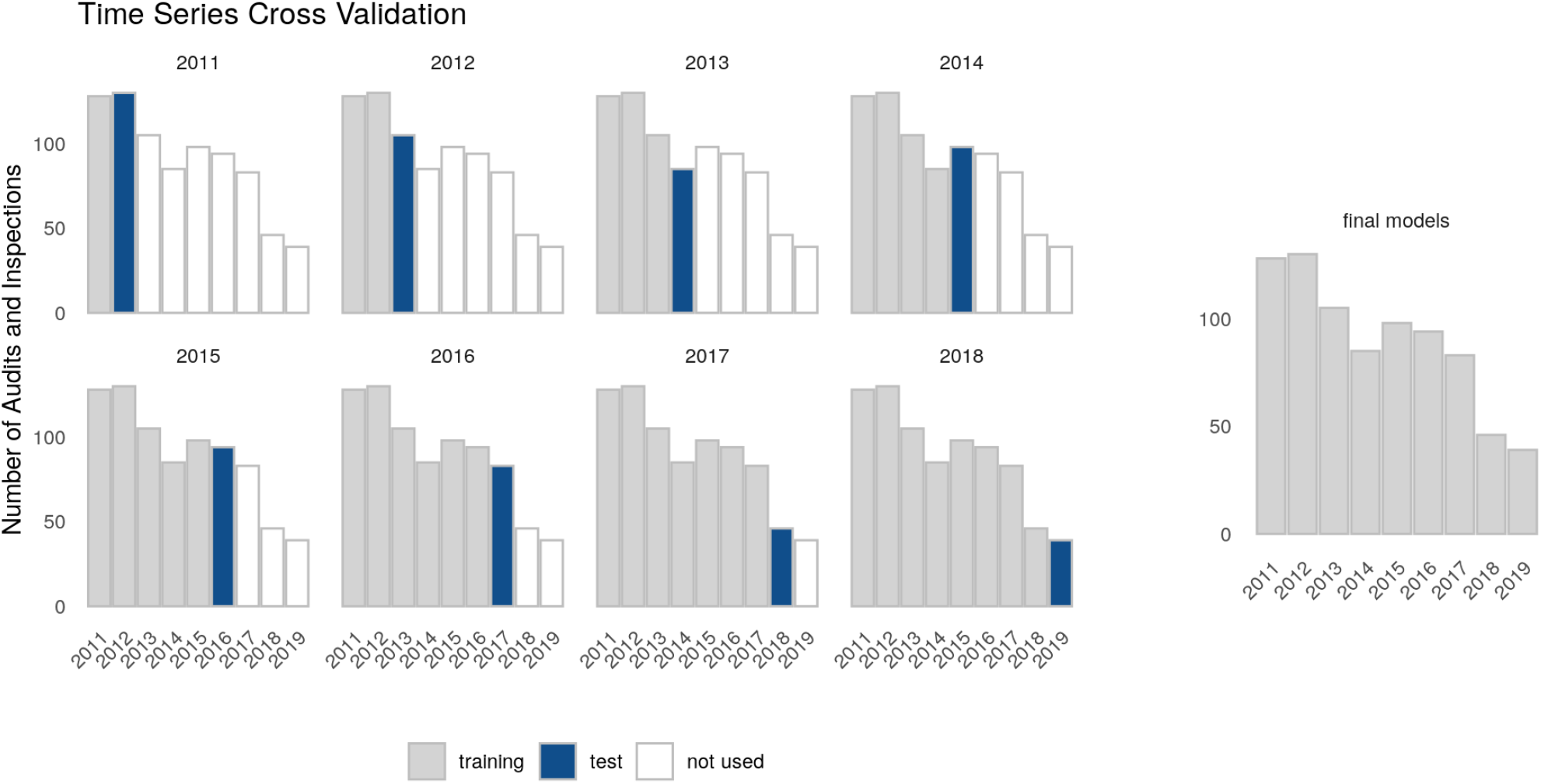
Time Series Cross Validation

As we were able to create more site-specific features, risk estimates for two sites from the same study were more likely to be different with more gradual increases and decreases. We therefore did not remove audits/inspection of previously audited/inspected studies from our test sets. The benefit of additional data points for calibration and performance estimates outweighs the risk of overestimating performance due to data leakage of study related attributes. As we expect risk estimates to change more gradually as site features change, a linear fit seemed more appropriate than the manually fitted step function we previously used [1].

## Results

### Performance and Model Characteristics

We were able to model audit and inspection finding risk for 4 out of 5 CIFs which could estimate differences in risk between 25 and 41% for different sites with AUCs between 0.59 and 0.63 and Brier Scores [6] from 0.19 to 0.25 (Table 1; Fig. 2). While classification performance as measured by AUC is poor, the calibration of the native risk predictions as measured by the Brier Score with 0 (perfect accuracy) to 1 (perfect inaccuracy) can be described as adequate. A predictive difference in risk by 25% or more between sites has clear business value and can influence business decisions. We were not able to model the impact factor sponsor oversight properly (Table 1; Fig. 2) most likely because we were lacking relevant features.

**Figure 2.**
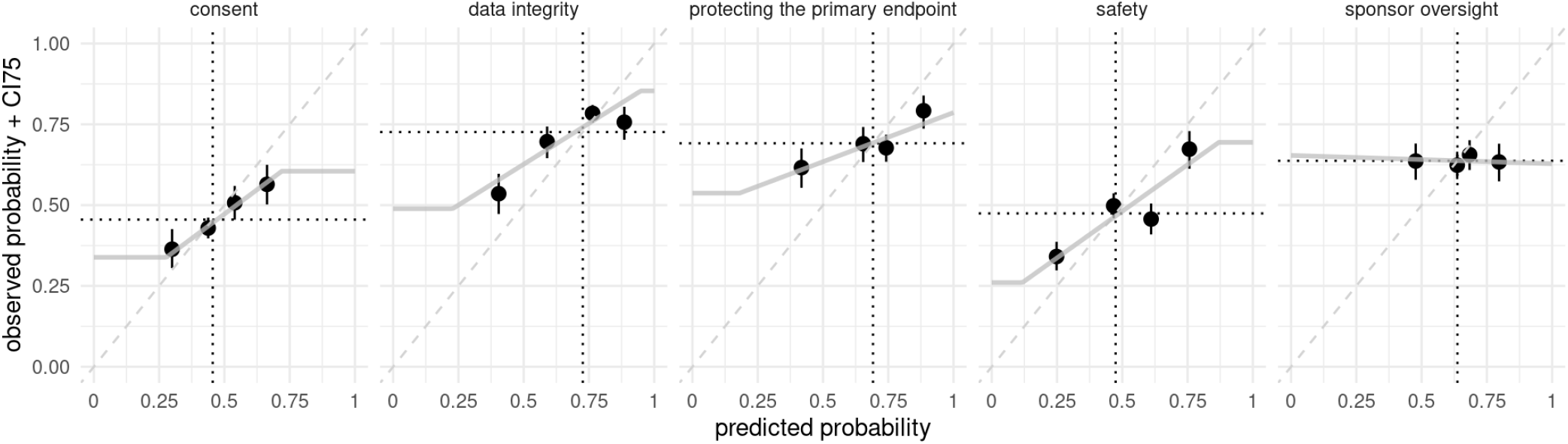
Calibration

### Features

The identified risk factors could be categorized into 7 groups (AE, Issues and Deviation, Data Queries, Geographical Populations, Parallel Trials and Study Characteristics; Table 2). AEs, data queries, and issues and deviations represented frequent on-site events that were connected to heavily regulated operational processes and left a coherent data trail. Thus, event frequencies and processing times could serve as proxy measures for operational quality and were likely to influence the risk of findings.

**Table 2.** Features Contributing to each CIF Model - Features Generally Correlate Positively with Audit Finding Risk Unless Indicated Otherwise (Green Downward Arrow

| Clinical Impact Factor |  |  |  |  |
| --- | --- | --- | --- | --- |
| Feature Group | Consent | Data Integrity | Protecting Endpoints | Safety |
| Issues and Deviations | ▲ # late issues<br>▲ # minor deviations per days on study | ▲ # overdue issues at time of audit | ▲ # late issues<br>▲ # minor deviations per days on study |  |
| Geographical Population<sup>1</sup> | ▲ total population | ▲ male to female ratio in 18-39 age group |  | ▲ % over 60 years |
| Patient AEs and Visits | ▲ % visits not done |  | ▲ AE reporting time | ▲ AE per visit<br>▲ SAE per visit |
| Data Queries<sup>2</sup> |  | ▲ open queries per days on study | ▲ query processing time |  |
| Parallel Trials<sup>3</sup> |  | ▲ active Roche trials in the past year in the same therapeutic area |  |  |
| Study Characteristics | ▲ pediatric |  |  | ▲ cancer |
|  |  |  |  | ▼ neuro-psychiatric |
|  |  |  |  | ▼ autoimmune |
| <sup>1</sup>Living Within a 100km Radius Around Site<br><sup>2</sup>Automatic or Manual Data Queries directed towards Site by Sponsor |  |  |  |  |

## Discussion

### Interpretation

We were able to identify 15 quality indicators - of which 13 were site specific - that could be linked with an increase or decrease in audit and inspection finding risk. The final model features were the result of a hypothesis-driven selection since we involved SMEs in the process. An algorithmic feature selection would have been unrealistic in this low signal-to-noise scenario which would have resulted in uninterpretable false-positive features. Although we could not conclude that there was a causal relationship between the model coefficients and the modeled events, many of them were process-related and would have naturally been identified by SMEs during process monitoring. This made them easy to integrate into a risk-based quality monitoring data product.

### Clinical Impact Factor: Consent (Fig. 3)

Informed consent requires paper signatures on forms that include the most up to date study information. This process is not yet fully digitized in most studies [7]. Therefore, a common auditing activity is to verify the paper signatures. If signatures were missing or were obtained too late a consent finding would be raised. The risk for such findings was increased for pediatric studies which require signatures from each parent. Moreover, when patients missed scheduled visits they might not have been able to reconsented in time. Consequently, risk increases with higher percentages of missed visits.

**Figure 3.**
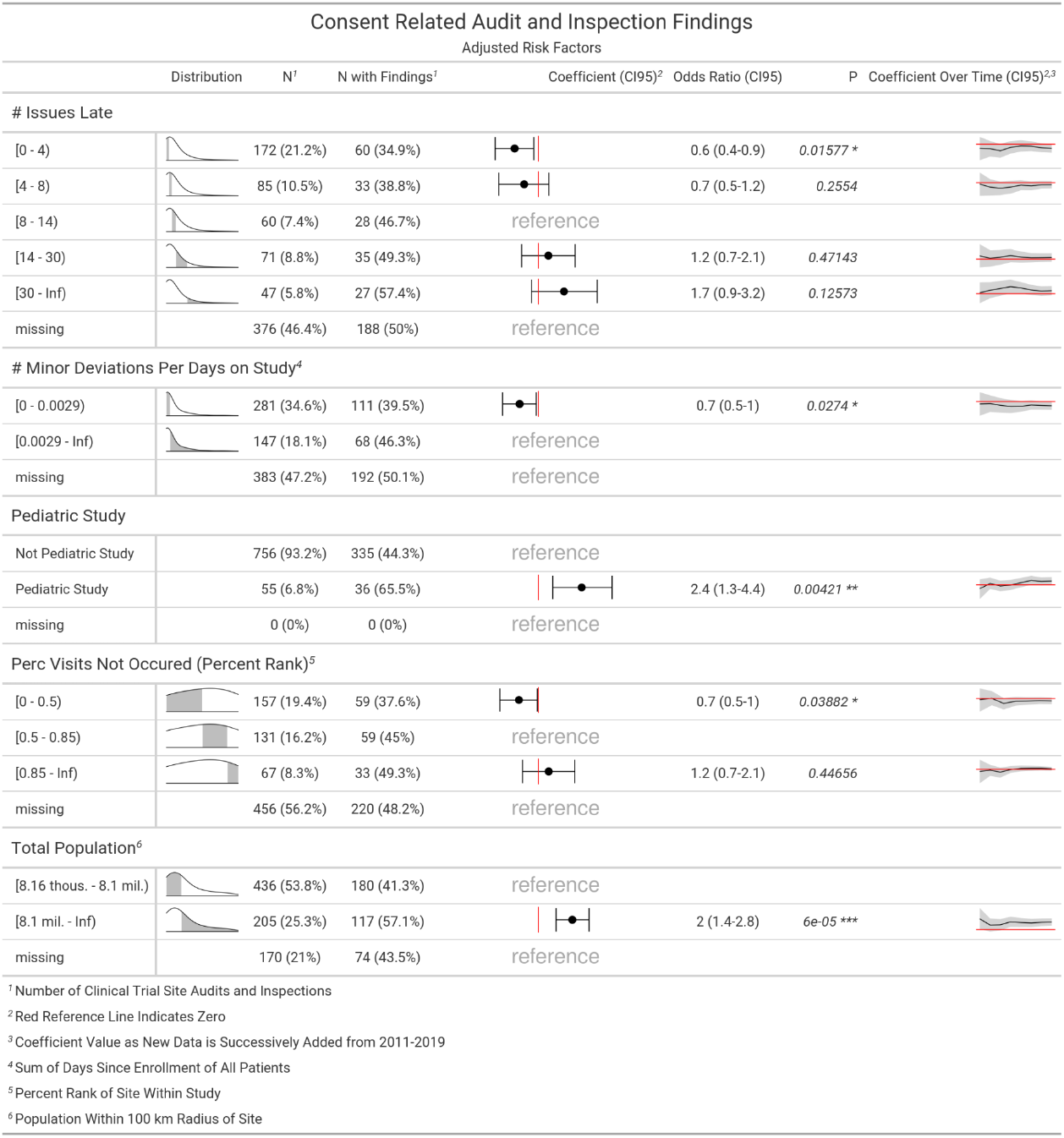
Clinical Impact Factor: Consent

Site issues and protocol deviations must be captured in an issue log and need to get resolved within a specific timeframe. Issues can be classified as protocol deviations that can be graded as minor or major. A high number of minor deviations overall, as well as a high number of late-resolved issues, could theoretically indicate general quality issues at a site. Both metrics increased consent finding risk as well as protecting the endpoints finding risk (Table 2 and Fig. 6). Of note, the overall issue and deviations generation rate is dependent on the number of patients processed by a site, so we needed to normalise in order to compare different sites. However, all sites should resolve their issues in time, so the absolute number of late issues carried similar weight for high and low enrolling sites. The absolute number of late issues was a better risk indicator than any normalised version. An increase in consent risk could also be found for sites located in densely populated areas. We could not see how this connected to the consent process. We did not find that population density was influencing any of the other risk models, thus we could only suspect that there was a strong unknown confounding variable that we were not yet capturing.

**Figure 4.**
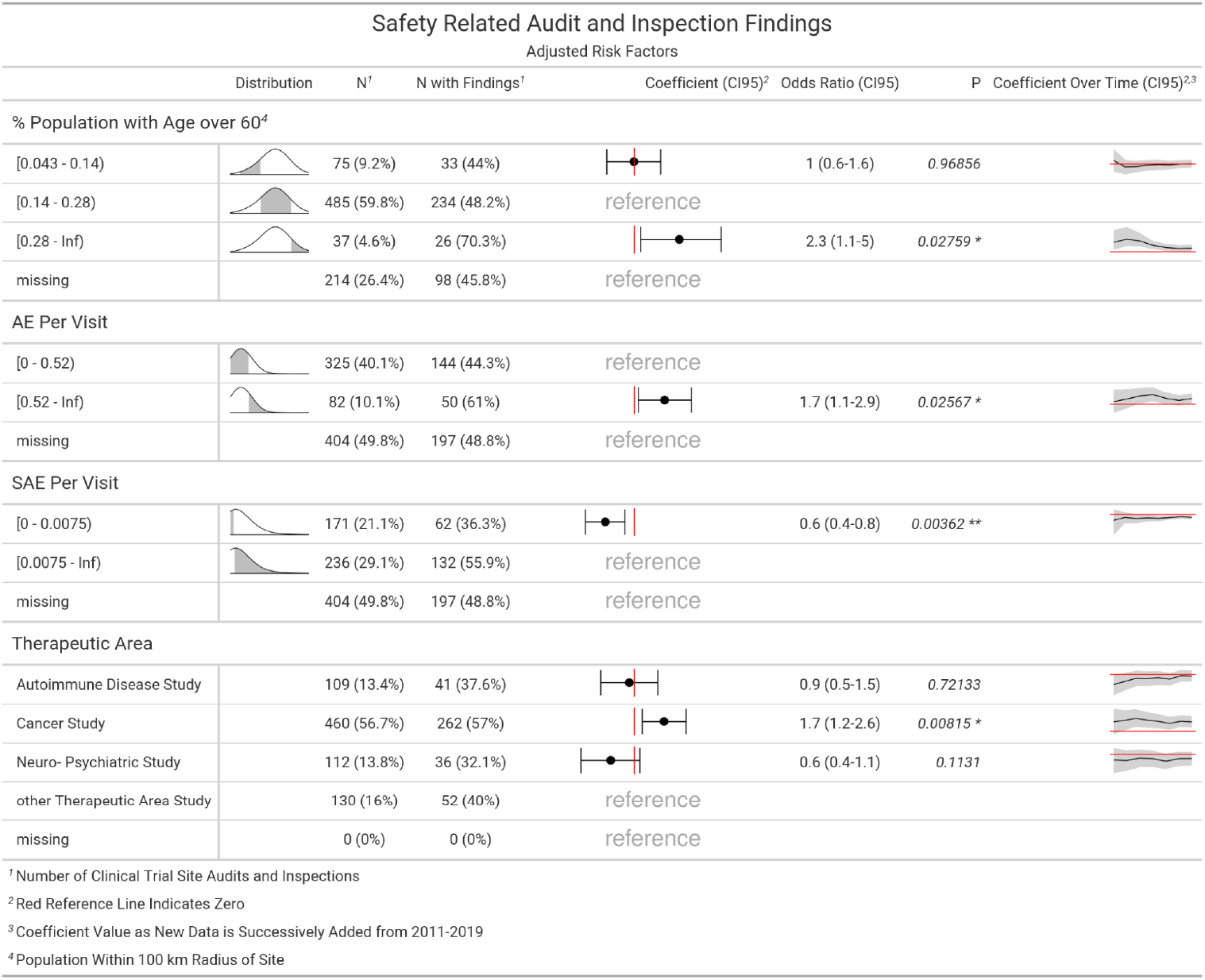
Clinical Impact Factor: Safety

**Figure 5.**
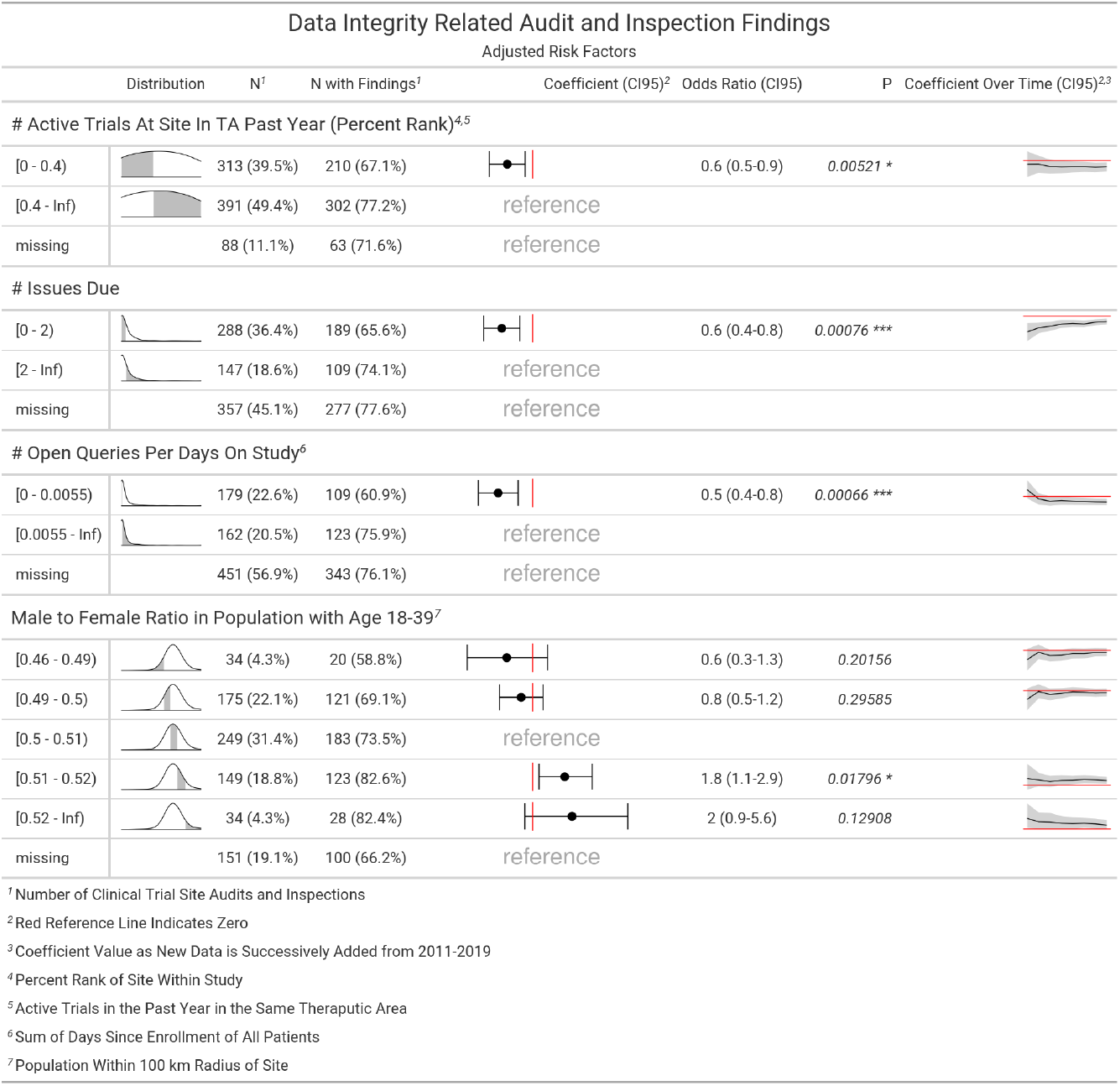
Clinical Impact Factor: Data Integrity

**Figure 6.**
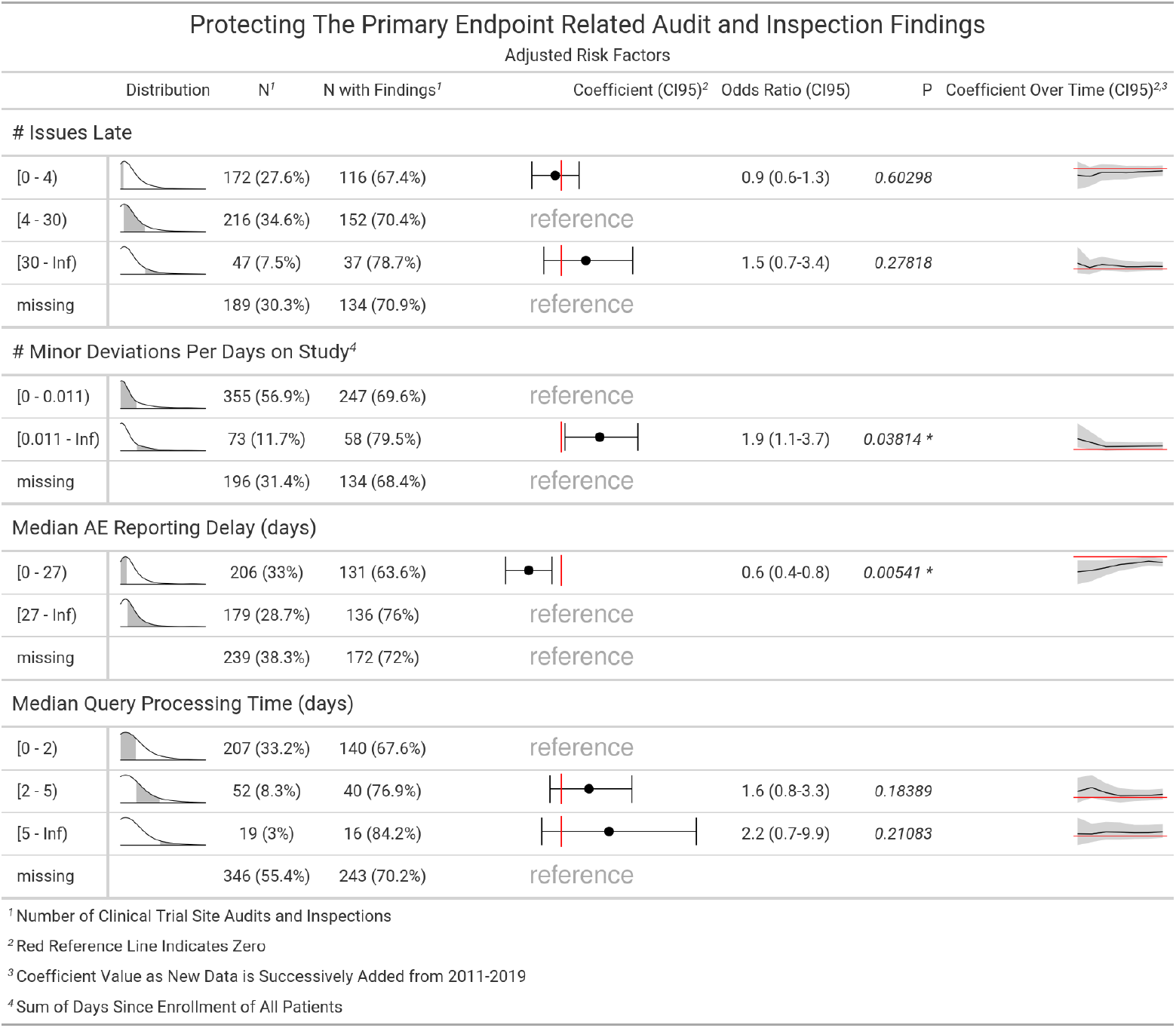
Clinical Impact Factor: Protecting the primary endpoint

### Clinical Impact Factor: Safety (Fig. 4)

AEs need to be adequately recorded into the medical database followed by a medical seriousness and causality assessment by qualified site staff. For serious AEs (SAEs) accelerated reporting timelines apply. Any detectable failure in this process would trigger safety findings. Accordingly, high rates of AEs increased the risk of safety findings while a low rate of SAEs or the absence of SAEs decreased risk. Independently of AE rates, cancer studies had an increased risk for safety findings as did sites that were located in a region with a high percentage of over 60-year-old in the geographical population around the site. We could speculate that older or terminally ill patients were more likely to suffer from concomitant diseases which added additional complexity to the adequate AE capture and causality assessment.

### Clinical Impact Factor: Data Integrity (Fig. 5)

Whenever there were mismatches between source data records and the clinical databases, a data integrity finding was raised. Invalid entries into the clinical database or a site monitor discovering questionable entries could trigger a data query that needed to be addressed by the site staff. In preparation for an audit or inspection, sites usually tend to double check data entries and resolve issues and open data queries. If the number of open queries and the number of overdue issues were low, risk for data integrity findings was decreased (possibly indicating that the site was well prepared for the audit/inspection). Furthermore, a comparatively low number of active trials in the last year at the site and the number of issues that were due at the time of the audit both decreased data integrity finding risk. These last two risk factors were not directly connected to data integrity and thus should be viewed as generalizable site quality indicators. Additionally, a higher ratio of females in the younger working population aged 18-39 would decrease the risk of data integrity findings, which geographically correlated with larger urban areas, which in turn had a higher concentration of general research centers. This was an interesting coincidental yet valuable correlation that we plan to investigate further.

### Clinical Impact Factor: Protecting the primary endpoints (Fig. 6)

Root causes for findings that were raised because the primary endpoints were at risk were numerous. Among the most frequent were inadequate study documents, mishandling of samples or the investigational medicinal product (IMP) and mismanaged protocol deviations. In this category we mostly identified risk factors that were indicative of overall site operational quality, some of which also influenced risk for findings for other CIFs such as number late issues, number of minor deviations and query processing time. Furthermore, a speedy reporting of AEs decreased risk as well which was potentially another operative site quality indicator.

## Challenges and Limitations

Using more operational site features over the more static study features of our previous iteration [1] was a major improvement. This resulted in more diverse risk predictions for all sites in a given trial with risk estimates that would continuously adjust as sites continued to participate in the trial creating new data points. However, none of the new operational features correlated with sponsor-oversight-related audit and inspection findings. Useful features could probably be engineered from monitoring visits, source data verification and vendor management, but we have not yet been able to obtain that data electronically for a sufficient fraction of previously audited sites. It was important to note that despite our effort to validate our models using time-series cross validation and calibration we have merely modeled the risk of historic investigator site audit and inspection findings. The COVID-19 pandemic has accelerated the decrease of traditionally conducted audits and inspections and remote quality activities are getting more and more common [8]. The number of traditional site audits in 2020 was already too low to include the data in this iteration. Sites audited in the past are often the sites that have recruited the most patients and thus carried the highest risk for the study outcome. Furthermore, findings were restricted to quality issues that could be identified by an auditing team on site. The risks and the quality indicators are integrated into a clinical analytics dashboard used by quality leads to manage audit focus and audit target selection. The risk as estimated by our models can only be used for a risk-based quality assurance strategy. It cannot be used to mitigate the individual risk of individual sites since we cannot assume a causal inference between the risk factors and the quality issues identified by the audits and inspections.

## Conclusion

As travel and physical access to sites is getting more restrictive, new complementing strategies for QA based on remote data analytics are emerging. Monitoring quality indicators and audit finding risk assessment could help to manage audit target selection and audit focus. To establish regulatory and industry trust, and to foster adoption of analytics-driven QA, we will continue to focus our effort on cross-company collaboration [9] and data sharing.

## Data Availability

Data sharing is not applicable to this article as no datasets were generated or analysed during the current study.

https://github.com/openpharma/quality_risk_assesment_clinical_trials

## Notes

**Conflict of Interest** Björn Koneswarakantha and Timothé Ménard were employed by Roche at the time this research was conducted.

### Competing Interest Statement

Björn Koneswarakantha and Timothé Ménard were employed by Roche at the time this research was conducted.

### Funding Statement

Development of the statistical model was funded by Roche.

### Summary of Updates

All sections of the manuscript have been revised for further clarifications on the study scope and objectives.

## References

1. Koneswarakantha, B., Ménard, T., Rolo, D. et al. Harnessing the Power of Quality Assurance Data: Can We Use Statistical Modeling for Quality Risk Assessment of Clinical Trials? Ther Innov Regul Sci 54, 1227–1235 (2020). https://doi.org/10.1007/s43441-020-00147-x

2. Zou, M., Barmaz, Y., Preovolos, M. et al. Using Statistical Modeling for Enhanced and Flexible Pharmacovigilance Audit Risk Assessment and Planning. Ther Innov Regul Sci 55, 190–196 (2021). https://doi.org/10.1007/s43441-020-00205-4

3. Yeo, IK., Johnson, RA. A new family of power transformations to improve normality or symmetry. Biometrika, 87(4):95 (2000). https://doi.org/10.1093/biomet/87.4.954

4. James, G., Witten, D., Hastie, T., Tibshirani, R. An introduction to statistical learning. 1st ed. New York: Springer (2013). https://doi.org/10.1007/978-1-4614-7138-7

5. Bergmeir, C., Benítez, JM. On the use of cross-validation for time series predictor evaluation. Information Sciences 191, 192–213. (2012) https://doi.org/10.1016/j.ins.2011.12.028

6. Brier, GW. Verification of forecasts expressed in terms of probability. Month Weather Rev. 1950;78(1):1–3. https://doi.org/10.1175/1520-0493(1950)078%3C0001:VOFEIT%3E2.0.CO;2

7. Bleiberg, H., Decoster, G., de Gramont, A., et al. A need to simplify informed consent documents in cancer clinical trials. A position paper of the ARCAD Group. Ann Oncol. 28(5):922–930 (2018). https://doi.org/10.1093/annonc/mdx050

8. Ménard, T., Bowling R., Mehta P., et al. Leveraging analytics to assure quality during the Covid-19 pandemic - The COVACTA clinical study example. Contemporary Clinical Trials Communications 20, 100662 (2020). https://doi.org/10.1016/j.conctc.2020.100662

9. Ménard, T., Young, K., Emerson, J., et al. Cross-company collaboration to leverage analytics for clinical quality and accelerate drug development: the IMPALA industry group. CPT Pharmacometrics Syst Pharmacol. 2021. https://doi.org/10.1002/psp4.12677

